# Mapping the national roll-out of social prescribing in England’s primary care system: an observational study of rates and patterns of referrals using data from the Clinical Practice Research Datalink (CPRD)

**DOI:** 10.1101/2025.04.04.25325237

**Authors:** Feifei Bu, Alexandra Burton, Naomi Launders, Amy E. Taylor, Alvin Richards-Belle, Stephanie Tierney, David Osborn, Daisy Fancourt

**Affiliations:** Department of Behavioural Science and Health, University College London; Wolfson Institute for Population Health, Centre for Psychiatry and Mental Health, Queen Mary University of London; Division of Psychiatry, University College London; Division of Surgery and Interventional Science, University College London; Nuffield Department of Primary Care Health Sciences, University of Oxford; North London NHS Foundation Trust

**Keywords:** social prescribing, non-clinical intervention, primary care, equality, equity

## Abstract

**Background:** Social prescribing (SP) is growing rapidly in England and across the world. However, whom it is reaching and how effectively it is being implemented remains unclear. This study aimed to assess longitudinal trends in SP in England’s primary care system, including growth trajectories and target alignment, sociodemographic profiles of referred patients, and predictors of service refusal over time.

**Methods:** This study analysed primary care records from 1.2 million patients from 1,736 practices in the Clinical Practice Research Datalink in England. We estimated SP trends between 2019 and 2023 using growth curve modelling on SP numbers at practice level. Descriptive analyses were used to show changes in sociodemographic profiles of SP patients over time. To assess sociodemographic disparities in service refusal (defined as having a medical code of ‘social prescribing declined’), we used multilevel logistic regression models stratified by year, accounting for nested data structure where patients were nested within practices.

**Findings:** As of the end of 2023, an estimated 9.4m GP consultations in England have involved SP codes, and 5.5m consultations have specifically led to SP referrals. In 2023, females constituted 60% of SP patients and ethnic minority groups represented 23%. Representation from patients living in more deprived areas increased from 23% to 42% between 2017-2023. Service refusal declined from 22% to 12% between 2019-2023. Age, sex and ethnicity were associated with service refusal across multiple years. In 2023, notably, all age groups had higher odds of refusal compared to the youngest age group. Females had 21% lower odds of refusal than males (95% CI=0.77-0.82), and patients from white ethnic backgrounds had 32% higher odds of refusal than ethnic minority patients (95% CI=1.26-1.39).

**Interpretation:** SP has expanded rapidly in England, far exceeding initial targets of 900,000 patients by 2023/24 and suggesting broad service acceptability. Progress is being made in reaching certain target groups such as more deprived communities. However, there are still disparities in accessibility and uptake, calling for targeted strategies to address underlying inequalities.

**Funding:** MQ Transforming Mental Health, Rosetrees-Stoneygate Trust Fellowship, National Academy for Social Prescribing

**Research in context:** *Evidence before this study:* We systematically searched PubMed, PsycINFO, Cochrane Library, Web of Science, and OpenGrey for studies (including grey literature) published in English between January 1980 and March 2025, using search terms such as ‘social prescribing’, ‘non-medical referral/intervention’, ‘non-clinical referral/intervention’, ‘community referral’, ‘referral scheme’. Prior evidence on rates and patterns of referrals was limited to small-scale evaluations, cross-sectional data, or regional analyses, with a lack of nationally representative longitudinal studies examining implementation trends or equity. Systematic reviews highlighted gaps in understanding disparities in service access and uptake. While the National Health Service (NHS) Long Term Plan (2019) set ambitious targets for SP, no studies had quantified progress toward these goals since the national rollout.

*Added value of this study:* Using primary care medical records from the Clinical Practice Research Datalink (CPRD), this study provides the first longitudinal, national analysis of SP implementation across 1.2 million patients in England. We demonstrate that SP referrals exceeded NHS targets by 27–51% in 2023, with at least 1.1–1.4 million patients receiving referrals. The analysis reveals significant progress in reaching deprived populations (representation increased from 23% to 42% between 2017-2023) but identifies persistent disparities in service uptake across age, sex, and deprivation groups.

*Implications of all the available evidence:* The rapid expansion of SP reflects its growing integration into primary care. However, persistent sociodemographic disparities highlight the need for targeted interventions to ensure equitable service access and uptake. This study provides policymakers with evidence to standardise referral protocols and allocate resources to underserved areas. Future research should rigorously track the implementation of SP, evaluate its long-term health outcomes and cost-effectiveness to fulfil its potentials as a key component of universal personalised care.

## Introduction

Social prescribing (SP) is an innovative approach to healthcare that connects patients to non-clinical services in their local community to support their health and wellbeing.^1^ These non-clinical services include a wide range of activities such as exercise, volunteering, arts and culture, counselling, befriending, training courses, housing support, benefits and employment advice.^2^ SP can be implemented through various models, tailored to the specific needs of communities and healthcare systems.^3^ In England, the predominant model is the general practitioner (GP)-link worker model where a healthcare professional in primary care (usually a GP) refers patients to a link worker (or other similar professionals) who then works with the patient to develop a personalised care plan that connects them to community support and interventions.^2^ The importance of SP lies in its potential to address the social, emotional, and practical needs of patients that are often interrelated to medical needs but are not covered by clinical treatments, and which form an estimated 20% of GP consultations.^4^ As such, SP provides a more holistic approach to patient care that complements existing clinical interventions, bridging critical service gaps. It also has the potential to alleviate pressure on primary care by diverting these non-clinical consultations to community services. As part of population health management strategy, SP may reduce health inequalities by improving service accessibility in deprived communities and actively engaging marginalised populations.^5^

In 2019, SP became an established part of the healthcare system in England with the publication of the National Health Service (NHS) Long Term Plan.^6^ This led to a roll-out of link workers across England with funding announced for 1,000 link workers in 2019. In 2022, it became a formal mandate for every Primary Care Network (a group of neighbouring GP practices^6^) to provide SP as part of its service,^7^ and in 2023, the NHS Long Term Workforce Plan included a rising commitment to fund 9,000 link workers by 2036/2037.^8^ However, there are concerns that SP may not effectively address health inequalities and could even exacerbating them by disproportionately benefiting less disadvantaged individuals.^9,10^ Analysis of the roll-out of link workers within Primary Care Networks has reported that the NHS did not meet its target of employing 1,000 link workers by 2020/21 and found inequalities geographically, with areas that recorded the greatest need for additional support recording the lowest levels of link worker employment across England.^11^ However, analysis of data from a large cloud-based SP referral-management platform, from over 160,000 patients who had received SP referrals, suggested the number of referrals in more deprived areas may be higher than in less deprived areas.^2^ This finding is echoed in self-reports of SP referrals from older adults in national cohort data, which again found that those receiving benefits and with lower wealth were more likely to report having received a referral.^12^ Nonetheless, we currently lack any large-scale analyses of how many people have been offered SP through the GP-link worker model and how equitable these referrals have been over time. NHS England set the target of referring over 900,000 people to SP by 2023/2024, but whether this has been met is undetermined.^6^

Drawing data from national primary care records, the aims of the present study were threefold: (1) to depict how SP activity in primary care has developed over the past 15 years in England, estimating the number of referrals to date compared to national targets, estimating SP frequency year on year, and tracking trajectories of growth; (2) to explore the sociodemographic profiles of adult patients referred to SP and their trends over time; (3) to map uptake vs refusal of SP and identify patterns and trends in predictors of SP service refusal. These aims are critical to understanding the growth and future potential of SP at a key moment in future planning for the NHS in England.

## Methods

### Data

Data were from the Clinical Practice Research Datalink (CPRD); a research data service that collects anonymised patient data routinely from a network of over 2000 GP practices across the UK. CPRD contains rich data on demographics, diagnoses and symptoms, prescriptions, tests and referrals from over 60 million patients over more than 35 years since 1989, including over 18 million currently registered patients. This study focused on England using data from CPRD Aurum which covers over 20% of general practices and has been shown to be representative of the English population in terms of geographic area, deprivation, urbanicity, age and sex.^13,14^

To map rates and trends in SP activity (aim 1), data were analysed aggregately using all data with a medical code related to SP. Although our primary analyses focused on SP, as a point of comparison, we additionally ran analyses using medical codes relating to other common non-clinical interventions that are similar or closely related to SP, as detailed below, to understand how changes in SP compare to changes in these other interventions over time.

To map sociodemographic profiles of people referred to SP (aim 2) and patterns and predictors of service refusal (aim 3), we used patient level data. To be included in these analyses, patients had to be aged 18 or over who were permanently registered and with a minimum follow-up of 12 months for data quality control purposes.

Data extraction was carried out in November 2024, restricted to the period of 2009-2023, starting from 10 years before the publication of the NHS Long Term Plan and excluding incomplete data in 2024. In total, this provided an analytical sample of around 1.2 million patients from 1,736 practices for the analysis for research aim 1, 0.9 million patients for research aim 2, and 0.7 million for research aim 3.

### Measures

Consultations related to SP were identified using the code lists in table 1, which included any discussion related to SP. For research aim 1, we additionally considered medical codes related to other non-clinical interventions to provide context for assessing SP. These included community navigator, health coach, health trainer, and health and wellbeing worker/coach (Table 1). For research aims 2 and 3, we focused on SP codes.

**Table 1.** Data extraction code lists for social prescribing and other non-clinical interventions.

Notes: <sup>†</sup> SNOMED CT stands for Systematized Nomenclature of Medicine Clinical Terms, which is a clinical terminology
| Group | SNOMED CT terms<sup>†</sup> | Read code terms |
| --- | --- | --- |
| **Social prescribing** | Referral to social prescribing service <sup>t</sup><br>Social prescribing offered<br>Seen by social prescribing link worker <sup>t</sup><br>Signposting to social prescribing service<br>Review of social prescribing plan<br>Social prescribing declined <sup>r</sup><br>Social prescribing case closed<br>Social prescribing plan completed<br>Social prescribing for mental health<br>Not suitable for social prescribing<br>Referral to social prescribing service from other agency <sup>t</sup> | Referral to social prescribing service <sup>t</sup><br>Social prescribing offered<br>Social prescribing declined <sup>r</sup><br>Social prescribing for mental health |
| **Community navigator** | Seen by community navigator <sup>t</sup><br>Referral to community navigator <sup>t</sup> | Seen by community navigator <sup>t</sup><br>Referral to community navigator <sup>t</sup> |
| **Health coach** | Health coach initiated encounter <sup>t</sup><br>Referred for health coaching <sup>t</sup><br>Seen by health coach <sup>t</sup><br>Patient initiated health coach encounter <sup>t</sup><br>Referral for health coaching <sup>t</sup> | Health coach initiated encounter <sup>t</sup><br>Referred for health coaching <sup>t</sup><br>Patient initiated health coach encounter <sup>t</sup><br>Seen by health coach <sup>t</sup> |
| **Health trainer** | Referral to health trainer declined <sup>r</sup><br>Referral to health trainer <sup>t</sup><br>Seen by health trainer <sup>t</sup><br>Signposting to health trainer | Referral to health trainer declined <sup>r</sup><br>Referral to health trainer <sup>t</sup><br>Seen by health trainer <sup>t</sup><br>Signposting to health trainer |
| **Health & wellbeing worker/coach** | Seen by health and wellbeing coach <sup>t</sup><br>Signposting to health and wellbeing worker |  |

coding system that health professionals used to recode patient data. England started to implement it in a phased approach from 2018 to replace read codes. <sup>r</sup>: terms used to identify refusal of social prescribing; <sup>t</sup>: terms used to identify uptake of social prescribing when a social prescribing referral to link worker or other similar professionals was made by a primary care healthcare professional

**Table 2.** Sample counts, estimates and predictions from linear growth curve model on social prescribing 2019-2023.

|  | year | N consultations<br>in CPRD | Predicted N consultations and<br>95% CI in England † | N patients<br>in CPRD | Predicted N patients and 95%<br>CI in England † | N practice<br>in CPRD | N practice<br>in England ‡ |
| --- | --- | --- | --- | --- | --- | --- | --- |
| All social<br>prescribing codes: | 2019 | 46311 | 267461 [143723, 391199] | 34962 | 235514 [152005, 319023] | 1666 | 6836 |
|  | 2020 | 315333 | 1093958 [995646, 1192271] | 195277 | 630711 [564462, 696961] | 1700 | 6616 |
|  | 2021 | 460471 | 1901655 [1780867, 2022443] | 246137 | 1018805 [937608, 1100001] | 1683 | 6522 |
|  | 2022 | 737616 | 2683114 [2513911, 2852318] | 389527 | 1394150 [1280459, 1507841] | 1668 | 6422 |
|  | 2023 | 874612 | 3433345 [3208050, 3658639] | 438625 | 1754262 [1602863, 1905661] | 1620 | 6311 |
|  | Total | 2434343 | 9379533 [8781650, 9977416] | -- | -- | -- | -- |
| Uptake codes<br>only: | 2019 | 22668 | 74048 [8349, 139747] | 18424 | 126967 [69560, 184374] | 1666 | 6836 |
|  | 2020 | 155929 | 611744 [555131, 668357] | 118512 | 420318 [373604, 467031] | 1700 | 6616 |
|  | 2021 | 273466 | 1135458 [1058034, 1212883] | 182197 | 707557 [648063, 767051] | 1683 | 6522 |
|  | 2022 | 441452 | 1642291 [1531899, 1752683] | 278257 | 985424 [901545, 1069303] | 1668 | 6422 |
|  | 2023 | 549190 | 2129087 [1982855, 2275318] | 309092 | 1252117 [1140641, 1363593] | 1620 | 6311 |
|  | Total | 1442705 | 5541636 [5160997, 5922276] | -- | -- | -- | -- |

Notes: † Estimates based on linear growth curve model (2019-2023) with random intercept and slope (see Table S1); ‡ Number of practices in December each year obtained from NHS digital, general practice workforce, 31 December 2024; -- Patient totals are not provided because the same patient may receive social prescribing across multiple years, summing annual counts would result in duplicate counting and artificially inflate the total number of patients.

For research aim 2, sociodemographic covariates included age groups derived from year and year of birth (18-29, 30-39, 40-49, 50-59, 60-69, 70-79, 80+), sex (male, female), ethnicity (white, ethnic minority), area deprivation in deciles measured by the index of multiple deprivation (IMD) based on patient postcode, area urbanicity (rural, urban) based on the Rural Urban Classification from the 2011 census and region of primary care practices (North East, North West, Yorkshire and the Humber, East Midlands, West Midlands, East of England, London, South East, South West).

For research aim 3, we were interested in medical codes indicating whether SP referrals were declined or accepted. Service refusal was defined as ‘social prescribing declined’ in contrast to ‘referral to SP services (from other agency)/seen by SP link worker’ (service uptake). If refusal and uptake codes cooccurred, it was coded as refusal. We considered other codes if they were used in conjunction with SP codes in the same consultation (e.g. ‘social prescribing offered’ + ‘referral to community navigator’ coded as uptake). See Table 1 for details. For these analyses, we excluded any codes without any clear indication of uptake (e.g. ‘social prescribing offered’, ‘signposting to social prescribing service’, ‘social prescribing for mental health’). Service refusal/uptake was measured at patient level. If a patient had multiple consultations in a given year, the most recent consultation was used.

### Statistical Analysis

For research aim 1, data were first analysed descriptively showing the number of GP consultations related to SP and other non-clinical interventions by year between 2009 and 2023. We then fitted a series of unconditional growth curve models, using GP practices as the level 2 units. This was to examine the average growth trajectories of SP and other similar non-clinical interventions, as well as their variations across GP practices. The time variable, year, was initially used as a discrete variable represented by dummy variables, allowing the shape of trajectories to be freely estimated. Subsequently, linear growth models were applied where appropriate to balance model parsimony and mitigate overfitting risks. Based on estimates of an average practice, we calculated national estimates by multiplying results by the total number of GP practices in England in each year.^15^ Growth curve analyses were restricted to 2017-2023 due to small sample sizes in earlier years.

For research aim 2, we carried out descriptive analyses showing the sociodemographic profiles of patients with a SP code by year in the period of 2017-2023.

For research aim 3, we used multilevel logistic regression models to examine how sociodemographic factors (age, sex, ethnicity, deprivation, urbanicity, region) were associated with service refusal, accounting for the nested data structure (patients within practices), which were stratified by year. These analyses were restricted to periods of 2019-2023 due to small sample sizes in earlier years.

Missing data were minimal for most sociodemographic covariates except for area deprivation and urbanicity (∼20%) that were obtained via postcode linkage. We used complete case analysis for descriptive analyses (aim 2), but multiple imputation (MI) with chained equations (MI=20), were carried out for regression analyses (aim 3) under the assumption of missing at random. Further, we conducted sensitivity analyses using delta adjustment to assess the robustness of the results under different scenarios of missing not at random. All analyses were conducted in Stata V18.

## Results

### Research aim1: Trends in SP over time

SP was implemented sparingly in England before 2019. It began to increase rapidly and steadily after the national roll-out in 2019, even during the COVID-19 pandemic (Figure S1a). This marked a clear change from prior to 2019, when there had only been small increases in similar non-clinical interventions, although it is notable that community navigator and health coaching had begun rising from 2015 (Figure S1b). Other than a slight decline in health trainers, these other non-clinical services have continued to grow, suggesting that SP did not simply replace existing non-clinical services.

Results from growth curve models with flexible trajectories suggested the rise in SP was approximately linear since 2019 (Figure 2a). After fitting a linear model, we estimated an annual increase by roughly 126 consultations or 61 patients per GP practice between 2019-2023(Table 2). In 2023 alone, there were an estimated 544 consultations (95% CI: 508-579) and 278 patients (95% CI: 254-302) related to SP per practice. If extrapolated to represent all GP practices in England, we estimated that roughly 1.6-1.9 million patients had SP codes, based on the total number of 6,311 GP practices in England in 2023.^16^ If restricting to patients with referral codes only, the estimates were 1.1-1.4 million. To extrapolate across years, this equates to an estimated 8.8-10 million consultations including any discussions of SP from 2019 to the end of 2023, 5.2-5.9 million of which had SP codes specifically indicating that referrals took place (Table 2).

Growth trajectories for comparison activities are shown in Figure 1b-1e, which were largely consistent with the descriptive analysis (Figure S1) and were much smaller in numbers compared to SP.

**Figure 1.**
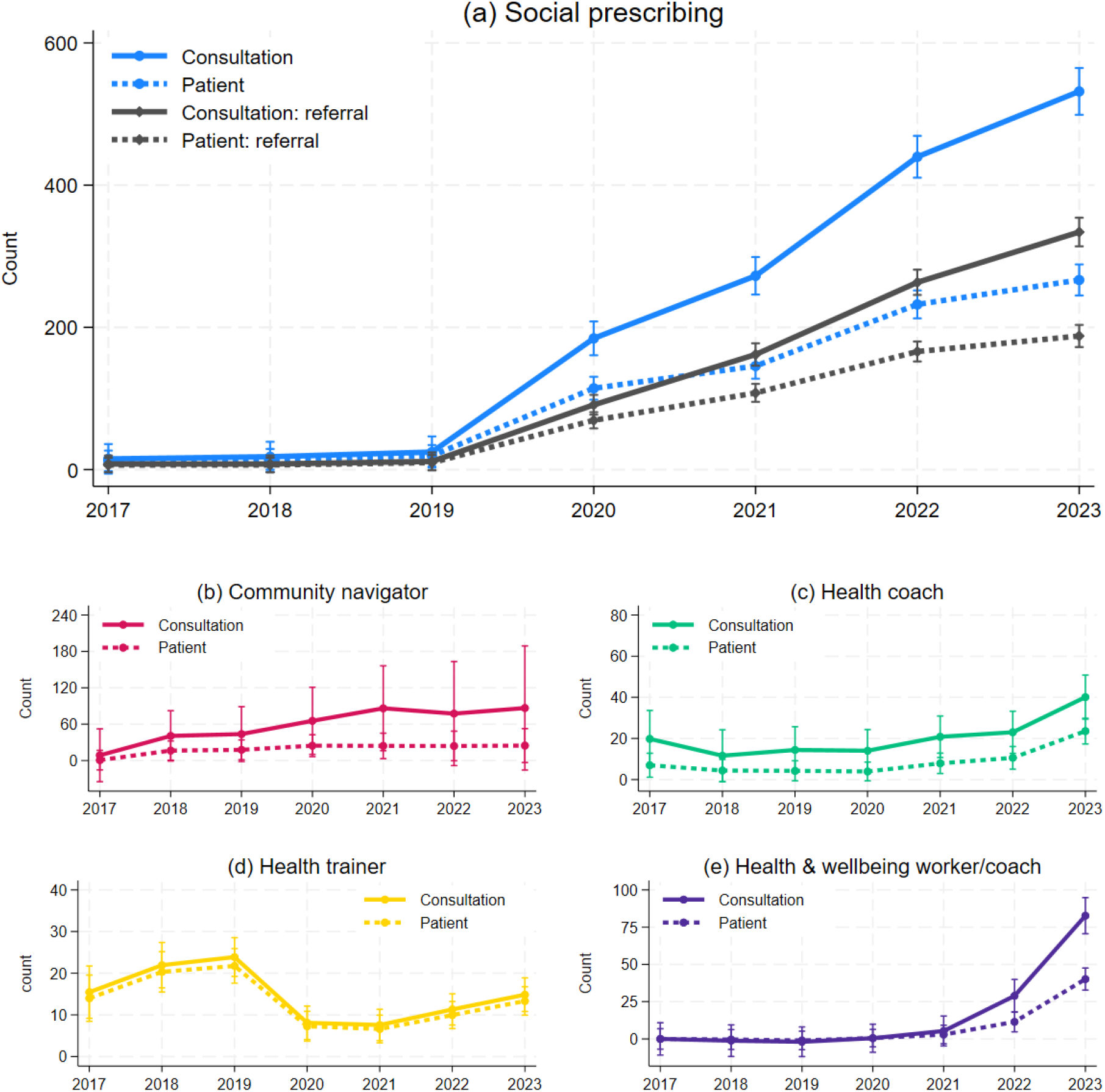
Average growth trajectories of numbers of consultations and patients across GP practices between 2017 and 2023 based on growth curve models with discrete time on the numbers of consultations and patients for social prescribing and other non-clinical interventions.

**Figure 2.**
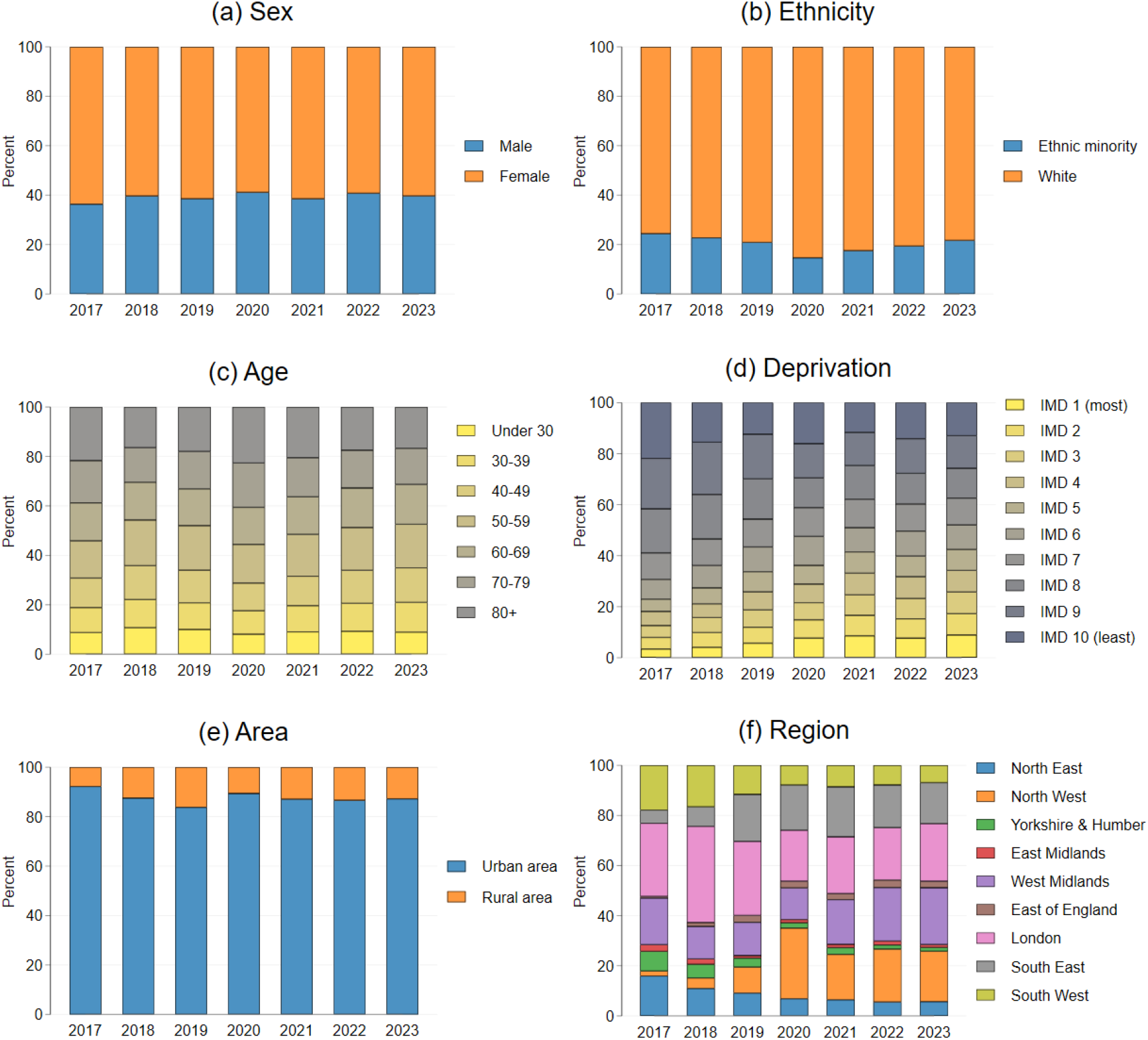
Changes in sociodemographic profiles of patients with social prescribing codes in CPRD Aurum (England, 2017-2023)

### Research aim 2: Sociodemographic patterning of SP referrals over time

In total, we identified around 0.9 million adult patients with SP codes between 2017 and 2023. Sample sizes varied across years and sociodemographic variables, ranging from 11260 to 720410 patients (Table S2-S5). Since 2017, females were overrepresented in SP consultations, accounting for around two thirds of patients consistently (Figure 3a, Table S2). Patients from ethnic minority backgrounds accounted for around 1 in 5 consultations in 2023, showing a gradual increase since 2020 (Figure 3b, Table S2). There was a slight but steady increase in the percentage of younger adults receiving SP codes since 2020 (Figure 3c, Table S4). Notably, patients from areas ranked in the top five most deprived deciles had also been increasing their access to SP, accounting for only 23% of referrals in 2017 but 42% in 2023 (Figure 3d, Table S4). Only around 13% of referrals were for individuals from rural areas, which had been stable since 2020 (Figure 3e, Table S2). Substantial changes in regional profiles are observable. For example, the percentage share of patients from London, the North East, Yorkshire and Humber, and South West decreased since 2017 as increasing numbers of patients from the West Midlands and North West had been accessing the service, now accounting for 23% and 20% respectively of all SP patients (Figure 3f, Table S5). However, it is important to note that caution should be taken comparing across regions as the distribution of GP practices within CPRD was not equal across geographical regions. Also of note is that the observed changes were unlikely driven by national sociodemographic shifts, given the stability of relevant characteristics during the study period (Table S6).

**Figure 3.**
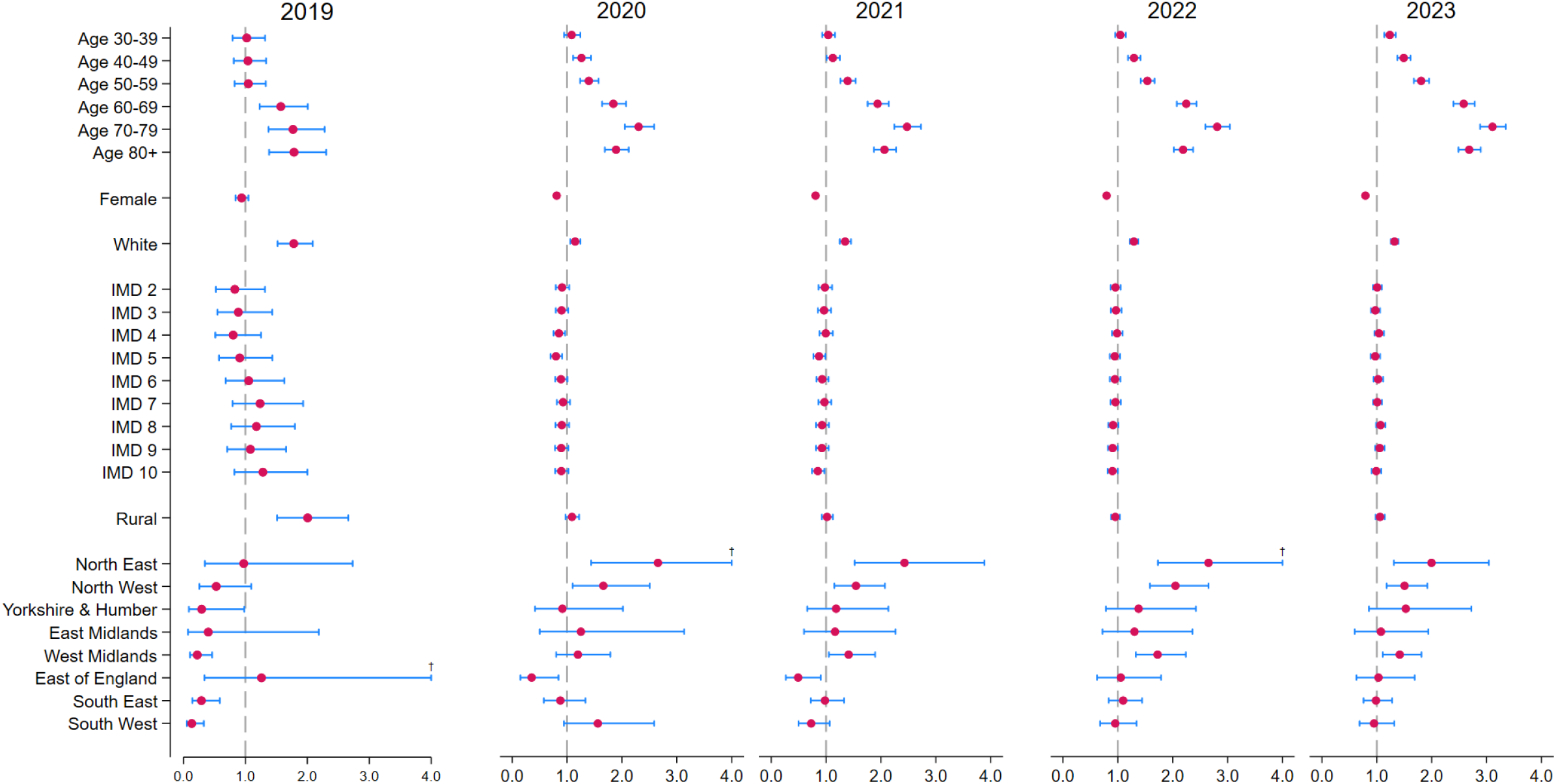
Odds ratios and their 95% confidence intervals from multilevel logistic regression models on service refusal fitted separately by year (MI=20) Notes: † large values capped at 4, reference categories are age 18-29, sex: male, ethnic minority, IMD 1-the most deprived, urban area, region: London.

### Research aim 3: Patterns and predictors of service refusal

In total, 0.7 million adult patents were included in the multilevel regression analyses between 2019 and 2023, with varying sample sizes and patient characteristics (Table S7). In 2019, 22% of patients had a record of service refusal, which declined to 11% in 2021 and remained stable since (Figure S2). It is of note that in 2019, most variance in service refusal for SP was between practices (72%), which dropped to 42% by 2023 (Figure S2). Results from regression models suggested that older age groups were more likely to refuse referrals, especially since 2020 (Figure 3, Table S8). While sex was not a predictor of refusal in 2019, females consistently had lower odds of refusal since 2020. In 2023, notably, females had 21% lower odds of refusal than males (95% CI=0.77-0.82). Patients of white ethnicity had higher odds of refusal than those from ethnic minority backgrounds in all years. There was no evidence that area deprivation or urbanicity was associated with SP refusal in any of the years, but there were some disparities across regions, including higher odds of refusal in the North East, West Midlands and North West compared to patients in London, since 2021. Results from models with sensitivity analyses using delta adjustment were largely consistent with the main analyses (Table S9).

## Discussion

Analysing data from CPRD, our study provides the first detailed analyses of the national roll-out of SP in England. Our findings demonstrate a rapid expansion of SP through GP practices since 2019. From 2019-2023, an estimated 8.8-10 million GP consultations involved discussions about SP, with over 5.2-5.9 million consultations leading to a referral. In 2023 alone, an estimated 1.6-1.9 million patients discussed SP with their GP and 1.1-1.4 million received a referral. These figures represent conservative estimates as they do not include other related codes (e.g. community navigators, health trainers, etc. unless used in combination with SP codes) that may (or may not) be used interchangeably with SP. They also do not include referrals from alternative routes, such as secondary care, voluntary and community sector organisations, statutory services, self-referral or patients who are referred but potentially not coded as such. These figures suggest SP has far exceeded the NHS Long Term Plan’s target of 900,000 patients by 2023/2024,^6^ underscoring a widespread recognition and adoption of SP within the primary care system.

While SP aims to reduce health inequalities, concerns have been raised that it may inadvertently exacerbate inequality by disproportionately benefiting less disadvantaged populations as a result of structural inequalities.^9,10^ In addition to concern about geographical inequalities in the roll-out of link workers, some pilot studies have reported lower referrals from individuals facing more socio-economic disadvantages.^11,17,18^ Our analyses found that in 2023, 42% of SP patients were from the top five most deprived deciles geographically. While equality or equity in SP distribution are not yet achieved, this figure is nearly double the 2017 percentage, with deprived-area representation increasing yearly. There have also previously been concerns about lower ethnic minority engagement in SP.^17–19^ But as of 2023, 21.7% of SP patients were ethnic minorities, surpassing the general population’s 19.3%.^20^ Furthermore, individuals from ethnic minority backgrounds appear more likely to take up SP referrals, after accounting for other factors. These findings show that SP has increasingly reached to traditionally underserved populations, marking significant progress toward accessibility equality. This aligns with national initiatives to reduce healthcare inequalities, such as the NHS Core20PLUS5 model lunched in 2021, targeting the most deprived 20% and locally identified high-risk groups, including ethnic minority communities.^21^

Despite these progresses, it is clear that work is still needed to ensure equity in SP. For example, we found an underrepresentation of rural residents comparing to national statistics (13% vs 17%),^22^ a gap persisting since 2020. This may be partially explained by the slightly overall underrepresentation of rural residents in CPRD (15% vs 17%),^14^ but it may be also related to limited community resources in some rural settings. In line with previous literature,^17,18^ our findings reveal a consistent overrepresentation of females among SP patients, despite an even sex distribution in the CPRD population.^13^ While this disparity may be partially attributable to higher consultation rates among females,^23^ sex differences in social, emotional or practical needs could also play a role. Further, females are more likely to take up referrals when offered, reinforcing sex-based disparities. Higher referral uptakes are also found among younger adults, who may benefit from greater acceptability of non-clinical interventions and fewer barriers in digital access and physical functioning. These findings corroborate earlier findings from the Social Prescribing Observatory,^24^ and highlight the importance of ensuring that existing accessibility disparities are not amplified through differences in service uptake by undertaking more extensive research to identify context-specific barriers across different SP stages of for underserved populations.^23^ Furthermore, although our data only focus on GP referrals, recent work using routine data covering a variety of referral pathways, suggests that young adults, individuals from more deprived groups and ethnic minority are more likely to access SP through non-clinical routes such as voluntary and community sector organisations, statutory services and self-referral.^2^ Therefore, it is important for future equity assessments to incorporate both GP and non-clinical pathways to avoid underestimating service penetration and to inform strategies for equitable referrals. Beyond individual sociodemographic factors, future research should also explore potential intersectional subgroups experiencing compounded disadvantages in SP referrals.

The main strength of our study is its use of a national longitudinal primary care dataset (CPRD) over a multiple-year period. Other strengths include the consideration of other related terms and robust analytical approaches accounting for nested data structure and missing data. However, several limitations should be noted. First, as our sample are restricted to SP patients, we cannot compare them to patients not considered for SP. Nevertheless, our study does provide valuable insights into the sociodemographic profiles of SP patients and their temporal trends. Second, while CPRD is generally recognised for its quality^26^, our study shares the limitation common to routine healthcare data regarding coding accuracy, particularly for SP, where both under-coding and over-coding may exist. CPRD data also currently lack granular detail in SP, with little information on referral reasons, prescribed interventions, patient outcomes and link worker characteristics (e.g. age, gender, work setting). But recent updates to coding systems, incorporating newly added SP codes, present opportunities to address these gaps in future work. Third, in this study, we have considered SP as well as related terms such as community navigator, health coach, health trainer, and health and wellbeing worker/coach. While these terms are occasionally used in combination with SP codes, they are predominantly recorded independently. Therefore, it is unclear whether they are used as precursors, subdomains, or standalone equivalents to SP. This lack of clarity highlights the urgent need for standardised definitions and coding practice to enhance data comparability for research and policy evaluation in this rapidly evolving field. Forth, our national projections of SP rely on the assumption that practices in CPRD constitute a nationally representative sample. While previous studies have established CPRD’s representativeness of the national population,^13,14^ its representativeness at the practice level is yet to be tested, meaning that these estimates should be interpreted with caution. Additionally, for research aim 3, we categorised SP codes intuitively based on whether they indicated acceptance or refusal. However, given the multiple codes available, it is possible that GPs may have used neutral codes (e.g. ‘social prescribing offered’) rather than acceptance/refusal specific codes, so we potentially underestimate the true samples for both acceptance and refusal. Finally, our analyses focus specifically on GP referrals, so they do not provide a complete picture of all SP referrals in England, and GP referrals should not be considered the only viable SP pathway.

Our study highlights the rapid expansion and evolving sociodemographic landscape of SP through GP practices, with referrals surpassing national targets by at least 27–51% in 2023. At a time when the future of the NHS is being redesigned, these results highlight the clear appetite for SP in primary care and the sizeable and growing number of patients who are being referred each year. Promisingly, there are increasing representations of ethnic minorities, younger adults, and those living in deprived areas accessing the service. However, addressing disparities in service accessibility and uptake requires targeted strategies and diverse referral pathways to ensure SP fulfils its promise. Moving forwards, it will be important to continue annual monitoring of the rates and patterns of SP referrals to ensure there are continued improvements in equity of access and support continuous quality improvement of the service. At local levels, GP practices are encouraged to consider barriers and enablers of access across the full pathway to reduce refusal rates and encourage sustained engagement. From a policy perspective, these findings also highlight a critical point: that the annual funding allocated to SP in the first four years alone has led to roughly 4-6 times the number of anticipated patient referrals. As such, SP is a much larger service within the NHS than previously acknowledged.

## Article information

### Contributors

FB, DF and AB conceptualised this study. AB and AT gained data access. ARB, AB and FB had accessed and verified the data. FB cleaned and analysed the data. FB and DF interpreted the results and wrote the manuscript. ARB, NL and DO advised on data management. All authors contributed to writing (review and editing). AB gained funding for the study. All authors approved the final manuscript and accepted the responsibility for the decision to submit it for publication.

## Acknowledgement

This work is supported by the MQ Transforming Mental Health and Rosetrees-Stoneygate Trust Fellowship (MQF22\6) awarded to AB and a grant from the National Academy for Social Prescribing awarded to DF. NL is supported by a Health Data Research UK personal fellowship. NL is affiliated to Health Data Research UK (Big Data for Complex Disease-HDR-23012), which is funded by the Medical Research Council (UKRI), the National Institute for Health Research, the British Heart Foundation, Cancer Research UK, the Economic and Social Research Council (UKRI), the Engineering and Physical Sciences Research Council (UKRI), Health and Care Research Wales, Chief Scientist Office of the Scottish Government Health and Social Care Directorates, and Health and Social Care Research and Development Division (Public Health Agency, Northern Ireland).

## Ethics statements

Ethical approval for this study was obtained from the Independent Scientific Advisory Committee of CPRD (protocol no. 24_004142).

## Patient and Public Involvement

Patients or the public were not involved in the design, or conduct, or reporting, or dissemination plans of our research

## Data availability statement

The original data from this study are provided by Clinical Practice Research DataLink (CPRD). Data cannot be shared publicly because they are not publicly available. CPRD may provide researchers with data following completion of their approvals and ethics process: https://www.cprd.com/research-applications

