## Supplement for "Mapping the national roll-out of social prescribing in England’s primary care system: an observational study of rates and patterns of referrals using data from the Clinical Practice Research Datalink (CPRD)"

Table S1 Results from linear growth curve model on numbers of social prescribing consultations and patients across practices (N=1,759)

| | All social prescribing codes | | | | | |
|---|---|---|---|---|---|---|
| | Consultations | | | Patients | | |
| | Coef. | 95% CI | p | Coef. | 95% CI | p |
| Time | 126.2 | [115.5, 136.9] | <0.001 | 60.9 | [53.7, 68.1] | <0.001 |
| Intercept | 39.1 | [21.0, 57.2] | <0.001 | 34.5 | [22.2, 46.7] | <0.001 |
| Var (Time) | 25589 | [23404, 27977] | -- | 11520 | [10535, 12597] | -- |
| Var (intercept) | 0.001 | [1.1e-74, 1.0e+68] | -- | 0.000 | [9.7e-132, 9.7e+117] | -- |
| Var (Residual) | 237248 | [229335, 245434] | -- | 108061 | [104458, 111789] | -- |
| | Uptake codes only | | | | | |
| | Consultations | | | Patients | | |
| | Coef. | 95% CI | p | Coef. | 95% CI | p |
| Time | 81.0 | [74.4, 87.5] | <0.001 | 45.0 | [39.8, 50.2] | <0.001 |
| Intercept | 10.6 | [1.0, 20.2] | <0.001 | 18.6 | [10.2, 27.0] | <0.001 |
| Var (Time) | 11945 | [11023,12945] | -- | 6540 | [6004,7125] | -- |
| Var (intercept) | 0.000 | [6.1e-105, 2.4e+93] | -- | 0.000 | [6.7e-99, 3.6e+87] | -- |
| Var (Residual) | 66564 | [64343, 68862] | -- | 51053 | [49351, 52815] | -- |

Table S2 Patient sociodemographic profiles by year: gender, ethnicity and area urbanicity

| | | Sex | | Ethnicity | | | Area | | |
|---|---|---|---|---|---|---|---|---|---|
| Year | N | Male | Female | N | White | Minority | N | Urban | Rural |
| 2017 | 14529 | 36.3% | 63.7% | 13687 | 75.6% | 24.4% | 11260 | 92.2% | 7.8% |
| 2018 | 21712 | 39.6% | 60.4% | 20031 | 77.3% | 22.7% | 18629 | 87.5% | 12.5% |
| 2019 | 32996 | 38.5% | 61.5% | 30700 | 79.1% | 20.9% | 27326 | 83.8% | 16.2% |
| 2020 | 242769 | 41.1% | 58.9% | 230422 | 85.4% | 14.6% | 204349 | 89.4% | 10.6% |
| 2021 | 354736 | 38.5% | 61.5% | 337880 | 82.5% | 17.5% | 297302 | 87.0% | 13.0% |
| 2022 | 588118 | 40.8% | 59.2% | 567476 | 80.6% | 19.4% | 471384 | 86.6% | 13.4% |
| 2023 | 720410 | 39.6% | 60.4% | 692131 | 78.3% | 21.7% | 568535 | 87.2% | 12.8% |

Table S3 Patient sociodemographic profiles by year: age group

| Year | N | Under 30 | 30-39 | 40-49 | 50-59 | 60-69 | 70-79 | 80+ |
|---|---|---|---|---|---|---|---|---|
| 2017 | 14529 | 8.8% | 8.8% | 8.8% | 8.8% | 8.8% | 8.8% | 8.8% |
| 2018 | 21712 | 10.7% | 10.7% | 10.7% | 10.7% | 10.7% | 10.7% | 10.7% |
| 2019 | 32996 | 10.0% | 10.0% | 10.0% | 10.0% | 10.0% | 10.0% | 10.0% |
| 2020 | 242769 | 8.1% | 8.1% | 8.1% | 8.1% | 8.1% | 8.1% | 8.1% |
| 2021 | 354736 | 9.0% | 9.0% | 9.0% | 9.0% | 9.0% | 9.0% | 9.0% |
| 2022 | 588118 | 9.3% | 9.3% | 9.3% | 9.3% | 9.3% | 9.3% | 9.3% |
| 2023 | 720410 | 8.9% | 8.9% | 8.9% | 8.9% | 8.9% | 8.9% | 8.9% |

Table S4 Patient sociodemographic profiles by year: area deprivation (1-most deprived, 10-least deprived)

| Year | N | IMD 1 | IMD 2 | IMD 3 | IMD 4 | IMD 5 | IMD 6 | IMD 7 | IMD 8 | IMD 9 | IMD 10 |
|---|---|---|---|---|---|---|---|---|---|---|---|
| 2017 | 11260 | 3.4% | 4.5% | 4.6% | 5.6% | 4.8% | 7.8% | 10.3% | 17.3% | 19.8% | 21.9% |
| 2018 | 18629 | 4.1% | 5.7% | 5.9% | 5.4% | 6.3% | 8.8% | 10.4% | 17.5% | 20.5% | 15.5% |
| 2019 | 27326 | 5.7% | 6.2% | 6.7% | 7.1% | 7.9% | 9.7% | 10.9% | 15.8% | 17.5% | 12.4% |
| 2020 | 204349 | 7.7% | 7.2% | 6.8% | 7.3% | 7.4% | 11.3% | 11.3% | 11.6% | 13.5% | 16.1% |
| 2021 | 297302 | 8.6% | 8.0% | 8.0% | 8.5% | 8.4% | 9.5% | 11.2% | 13.2% | 13.0% | 11.6% |
| 2022 | 471384 | 7.7% | 7.5% | 8.0% | 8.5% | 8.1% | 9.8% | 10.6% | 12.0% | 13.6% | 14.1% |
| 2023 | 568535 | 8.9% | 8.3% | 8.5% | 8.5% | 8.2% | 9.6% | 10.5% | 11.7% | 12.8% | 12.9% |

Table S5 Patient sociodemographic profiles by year: region

| Year | N | North East | North West | York & Humbe | East Mid | West Mid | East England | London | South East | South West |
|---|---|---|---|---|---|---|---|---|---|---|
| 2017 | 14529 | 15.9% | 2.0% | 7.8% | 2.7% | 18.6% | 0.6% | 29.2% | 5.3% | 17.8% |
| 2018 | 21712 | 11.0% | 4.2% | 5.5% | 2.1% | 12.9% | 1.5% | 38.4% | 7.7% | 16.6% |
| 2019 | 32996 | 9.1% | 10.3% | 3.6% | 1.0% | 13.3% | 2.7% | 29.5% | 18.7% | 11.6% |
| 2020 | 242769 | 6.9% | 28.2% | 2.1% | 1.3% | 12.6% | 2.7% | 20.4% | 18.0% | 7.9% |
| 2021 | 354736 | 6.4% | 18.1% | 2.7% | 1.3% | 17.8% | 2.4% | 22.7% | 19.9% | 8.6% |
| 2022 | 588118 | 5.5% | 21.1% | 1.6% | 1.6% | 21.3% | 3.0% | 20.9% | 17.0% | 7.9% |
| 2023 | 720410 | 5.7% | 20.1% | 1.5% | 1.1% | 22.7% | 2.6% | 23.0% | 16.4% | 6.9% |

Table S6 Sociodemographic composition of the national population 2017-2023

| | 2017 | 2018 | 2019 | 2020 | 2021 | 2022 | 2023 |
|---|---|---|---|---|---|---|---|
| Males | 49.0% | 49.0% | 49.0% | 49.0% | 49.0% | 49.1% | 49.1% |
| Females | 51.0% | 51.0% | 51.0% | 51.0% | 51.0% | 50.9% | 50.9% |
| Age: 18-29 | 19.8% | 19.6% | 19.3% | 19.0% | 18.7% | 18.7% | 18.8% |
| Age: 30-39 | 17.1% | 17.2% | 17.3% | 17.3% | 17.3% | 17.4% | 17.4% |
| Age: 40-49 | 16.8% | 16.5% | 16.3% | 16.2% | 16.0% | 15.8% | 15.8% |
| Age: 50-59 | 16.9% | 17.0% | 17.1% | 17.2% | 17.3% | 17.1% | 16.8% |
| Age: 60-69 | 13.3% | 13.2% | 13.2% | 13.3% | 13.5% | 13.7% | 13.9% |
| Age: 70-79 | 10.0% | 10.3% | 10.5% | 10.7% | 10.9% | 10.9% | 10.9% |
| Age: 80+ | 6.1% | 6.2% | 6.3% | 6.3% | 6.3% | 6.3% | 6.4% |
| Ethnicity: white | 83.9% | 83.6% | 83.2% | 82.9% | 81.7% | 82.5% | -- |
| Ethnicity: other | 16.1% | 16.4% | 16.8% | 17.1% | 18.3% | 17.5% | -- |
| Rural | 16.4% | 16.4% | 16.4% | 16.5% | 16.6% | 16.6% | -- |
| Urban | 83.6% | 83.6% | 83.6% | 83.5% | 83.4% | 83.4% | -- |
| IMD 1 | 20.0% | 20.0% | 20.0% | 20.0% | 20.0% | -- | -- |
| IMD 2 | 20.6% | 20.6% | 20.6% | 20.6% | 20.6% | -- | -- |
| IMD 3 | 20.3% | 20.3% | 20.3% | 20.3% | 20.3% | -- | -- |
| IMD 4 | 19.7% | 19.7% | 19.8% | 19.8% | 19.8% | -- | -- |
| IMD 5 | 19.4% | 19.4% | 19.4% | 19.4% | 19.4% | -- | -- |
| East Midlands | 8.6% | 8.6% | 8.6% | 8.6% | 8.6% | 8.6% | -- |
| East of England | 11.2% | 11.2% | 11.2% | 11.2% | 11.2% | 11.2% | -- |
| London | 15.8% | 15.8% | 15.8% | 15.7% | 15.6% | 15.5% | -- |
| North East | 4.7% | 4.7% | 4.7% | 4.7% | 4.7% | 4.7% | -- |
| North West | 13.1% | 13.1% | 13.1% | 13.1% | 13.1% | 13.2% | -- |
| South East | 16.4% | 16.4% | 16.4% | 16.4% | 16.4% | 16.4% | -- |
| South West | 10.0% | 10.0% | 10.0% | 10.1% | 10.1% | 10.1% | -- |
| West Midlands | 10.5% | 10.5% | 10.5% | 10.5% | 10.5% | 10.5% | -- |
| Yorkshire and The Humber | 9.8% | 9.7% | 9.7% | 9.7% | 9.7% | 9.7% | -- |

Notes: Data were obtained from the Office for National Statistics; -- No data available at the time of writing

Table S7 Sample characteristics in the multilevel logistic regression models by year

| | 2019 (n=13,686) | 2020 (n=82,209) | 2021 (n=119,886) | 2022 (n=185,850) | 2023 (n=206,416) |
|---|---|---|---|---|---|
| Age: 18-29 | 8.4% | 5.7% | 8.4% | 9.0% | 7.5% |
| Age: 30-39 | 11.3% | 7.3% | 10.3% | 11.2% | 10.6% |
| Age: 40-49 | 13.3% | 8.9% | 11.6% | 12.8% | 13.1% |
| Age: 50-59 | 17.9% | 13.8% | 15.9% | 16.8% | 17.3% |
| Age: 60-69 | 15.1% | 15.3% | 15.0% | 16.1% | 16.9% |
| Age: 70-79 | 15.2% | 22.1% | 17.1% | 16.0% | 16.3% |
| Age: 80+ | 18.9% | 26.8% | 21.7% | 18.1% | 18.4% |
| Sex: male | 38.2% | 41.2% | 39.2% | 39.4% | 39.8% |
| Sex: female | 61.8% | 58.8% | 60.8% | 60.6% | 60.2% |
| Ethnicity: white | 79.2% | 83.9% | 82.0% | 79.7% | 80.9% |
| Ethnicity: minority | 20.8% | 16.1% | 18.0% | 20.3% | 19.1% |
| IMD 1: most deprived | 5.2% | 9.0% | 8.5% | 8.2% | 11.7% |
| IMD 2 | 6.5% | 8.4% | 8.3% | 7.9% | 9.8% |
| IMD 3 | 6.1% | 7.9% | 8.7% | 8.1% | 10.0% |
| IMD 4 | 6.7% | 8.6% | 9.1% | 8.4% | 8.1% |
| IMD 5 | 7.2% | 8.4% | 8.4% | 7.9% | 7.9% |
| IMD 6 | 9.5% | 9.9% | 9.6% | 10.1% | 9.1% |
| IMD 7 | 10.5% | 10.6% | 10.9% | 10.5% | 10.0% |
| IMD 8 | 15.6% | 12.8% | 12.3% | 11.6% | 10.7% |
| IMD 9 | 18.7% | 12.9% | 12.7% | 13.4% | 11.7% |
| IMD 10: least deprived | 14.0% | 11.4% | 11.4% | 13.9% | 10.9% |
| Area: urban | 82.9% | 87.0% | 85.2% | 85.6% | 85.6% |
| Area: rural | 17.1% | 13.0% | 14.8% | 14.4% | 14.4% |
| Region: North East | 10.1% | 8.2% | 5.9% | 5.0% | 4.1% |
| Region: North West | 10.9% | 17.5% | 18.1% | 22.1% | 26.1% |
| Region: Yorkshire & Humber | 3.1% | 2.6% | 2.8% | 2.0% | 1.9% |
| Region: East Midlands | 1.1% | 0.8% | 1.0% | 1.9% | 1.1% |
| Region: West Midlands | 14.5% | 14.5% | 14.0% | 16.4% | 15.0% |
| Region: East of England | 3.7% | 4.7% | 3.8% | 4.6% | 4.0% |
| Region: London | 31.4% | 22.9% | 23.8% | 22.6% | 23.2% |
| Region: South East | 12.6% | 17.5% | 20.2% | 16.2% | 16.0% |
| Region: South West | 12.5% | 11.4% | 10.4% | 9.2% | 8.6% |

Table S8 Results on service decline from multilevel logit models by year using multiple imputation (MI=20)

| | 2019 Declined: 22.0% (n=17,493 N=857) | | | 2020 Declined: 18.5% (n=107,344 N=1,557) | | | 2021 (Declined: 10.7%) (n=150,399 N=1,643) | | | 2022 (Declined: 11.2%) (n=235,741 N=1,656) | | | 2023 (Declined: 11.2%) (n=266,193 N=1,616) | | |
|---|---|---|---|---|---|---|---|---|---|---|---|---|---|---|---|
| | OR | 95% CI | p | OR | 95% CI | p | OR | 95% CI | p | OR | 95% CI | p | OR | 95% CI | p |
| Age: 30-39 (vs <30) | 1.02 | [0.79,1.32] | 0.86 | 1.08 | [0.95,1.24] | 0.25 | 1.04 | [0.93,1.16] | 0.50 | 1.05 | [0.95,1.15] | 0.34 | 1.24 | [1.14,1.35] | <0.001 |
| Age: 40-49 (vs <30) | 1.04 | [0.81,1.34] | 0.75 | 1.26 | [1.11,1.44] | <0.001 | 1.12 | [1.01,1.25] | 0.04 | 1.29 | [1.19,1.41] | <0.001 | 1.49 | [1.37,1.61] | <0.001 |
| Age: 50-59 (vs <30) | 1.05 | [0.83,1.33] | 0.70 | 1.40 | [1.24,1.57] | <0.001 | 1.39 | [1.26,1.54] | <0.001 | 1.54 | [1.42,1.67] | <0.001 | 1.81 | [1.68,1.95] | <0.001 |
| Age: 60-69 (vs <30) | 1.57 | [1.23,2.01] | <0.001 | 1.84 | [1.64,2.07] | <0.001 | 1.94 | [1.76,2.14] | <0.001 | 2.25 | [2.07,2.43] | <0.001 | 2.58 | [2.40,2.78] | <0.001 |
| Age: 70-79 (vs <30) | 1.77 | [1.37,2.28] | <0.001 | 2.30 | [2.05,2.59] | <0.001 | 2.47 | [2.24,2.73] | <0.001 | 2.81 | [2.60,3.04] | <0.001 | 3.11 | [2.88,3.35] | <0.001 |
| Age: 80+ (vs <30) | 1.79 | [1.38,2.31] | <0.001 | 1.89 | [1.69,2.12] | <0.001 | 2.06 | [1.87,2.28] | <0.001 | 2.19 | [2.02,2.37] | <0.001 | 2.68 | [2.49,2.89] | <0.001 |
| Female (vs male) | 0.94 | [0.84,1.05] | 0.27 | 0.81 | [0.78,0.85] | <0.001 | 0.81 | [0.77,0.84] | <0.001 | 0.79 | [0.77,0.82] | <0.001 | 0.79 | [0.77,0.82] | <0.001 |
| White (vs other) | 1.78 | [1.52,2.09] | <0.001 | 1.15 | [1.06,1.24] | 0.001 | 1.35 | [1.25,1.45] | <0.001 | 1.29 | [1.22,1.37] | <0.001 | 1.32 | [1.26,1.39] | <0.001 |
| IMD 2 (vs 1) | 0.83 | [0.52,1.32] | 0.43 | 0.91 | [0.79,1.04] | 0.16 | 0.98 | [0.86,1.11] | 0.73 | 0.96 | [0.87,1.05] | 0.35 | 1.01 | [0.93,1.09] | 0.88 |
| IMD 3 (vs 1) | 0.89 | [0.55,1.43] | 0.62 | 0.90 | [0.79,1.02] | 0.10 | 0.96 | [0.85,1.09] | 0.55 | 0.97 | [0.87,1.07] | 0.50 | 0.97 | [0.89,1.06] | 0.52 |
| IMD 4 (vs 1) | 0.80 | [0.52,1.25] | 0.33 | 0.85 | [0.75,0.96] | 0.01 | 0.99 | [0.88,1.12] | 0.92 | 0.99 | [0.89,1.09] | 0.79 | 1.04 | [0.96,1.13] | 0.37 |
| IMD 5 (vs 1) | 0.91 | [0.58,1.44] | 0.68 | 0.80 | [0.70,0.91] | 0.00 | 0.87 | [0.77,0.99] | 0.03 | 0.94 | [0.86,1.04] | 0.25 | 0.97 | [0.89,1.06] | 0.50 |
| IMD 6 (vs 1) | 1.05 | [0.68,1.63] | 0.81 | 0.89 | [0.78,1.01] | 0.07 | 0.93 | [0.82,1.05] | 0.22 | 0.95 | [0.86,1.05] | 0.29 | 1.02 | [0.94,1.11] | 0.59 |
| IMD 7 (vs 1) | 1.24 | [0.79,1.93] | 0.35 | 0.93 | [0.81,1.05] | 0.25 | 0.97 | [0.86,1.09] | 0.62 | 0.96 | [0.87,1.05] | 0.37 | 1.01 | [0.93,1.09] | 0.84 |
| IMD 8 (vs 1) | 1.18 | [0.77,1.80] | 0.45 | 0.90 | [0.79,1.04] | 0.15 | 0.93 | [0.81,1.05] | 0.24 | 0.91 | [0.83,1.01] | 0.08 | 1.07 | [0.99,1.16] | 0.11 |
| IMD 9 (vs 1) | 1.08 | [0.71,1.66] | 0.72 | 0.89 | [0.78,1.02] | 0.10 | 0.92 | [0.81,1.05] | 0.21 | 0.91 | [0.82,1.00] | 0.05 | 1.05 | [0.97,1.14] | 0.22 |
| IMD 10 (vs 1) | 1.28 | [0.82,2.00] | 0.27 | 0.90 | [0.78,1.02] | 0.11 | 0.85 | [0.74,0.97] | 0.02 | 0.90 | [0.82,1.00] | 0.05 | 0.99 | [0.91,1.08] | 0.81 |
| Rural (vs urban) | 2.01 | [1.51,2.66] | <0.001 | 1.09 | [0.97,1.22] | 0.14 | 1.02 | [0.92,1.12] | 0.73 | 0.95 | [0.88,1.04] | 0.28 | 1.06 | [0.98,1.14] | 0.16 |
| North East† | 0.97 | [0.35,2.73] | 0.96 | 2.66 | [1.44,4.90] | 0.002 | 2.43 | [1.52,3.88] | <0.001 | 2.65 | [1.73,4.07] | <0.001 | 2.00 | [1.31,3.04] | 0.001 |
| North West† | 0.53 | [0.26,1.09] | 0.09 | 1.66 | [1.10,2.51] | 0.02 | 1.54 | [1.15,2.07] | 0.004 | 2.05 | [1.59,2.65] | <0.001 | 1.50 | [1.18,1.92] | 0.001 |
| Yorkshire & Humber† | 0.29 | [0.09,0.98] | 0.05 | 0.92 | [0.42,2.02] | 0.83 | 1.18 | [0.66,2.13] | 0.58 | 1.38 | [0.78,2.42] | 0.27 | 1.53 | [0.86,2.72] | 0.15 |
| East Midlands† | 0.40 | [0.07,2.19] | 0.29 | 1.25 | [0.50,3.13] | 0.63 | 1.16 | [0.60,2.27] | 0.66 | 1.30 | [0.72,2.36] | 0.38 | 1.08 | [0.60,1.94] | 0.81 |
| West Midlands† | 0.22 | [0.11,0.46] | <0.001 | 1.20 | [0.80,1.79] | 0.38 | 1.41 | [1.05,1.89] | 0.02 | 1.73 | [1.33,2.24] | <0.001 | 1.42 | [1.11,1.81] | 0.005 |
| East of England† | 1.26 | [0.34,4.68] | 0.73 | 0.36 | [0.15,0.84] | 0.02 | 0.49 | [0.27,0.90] | 0.02 | 1.05 | [0.62,1.79] | 0.85 | 1.03 | [0.63,1.69] | 0.91 |
| South East† | 0.29 | [0.14,0.59] | 0.001 | 0.88 | [0.58,1.34] | 0.54 | 0.98 | [0.72,1.33] | 0.89 | 1.10 | [0.83,1.44] | 0.51 | 0.98 | [0.76,1.28] | 0.91 |
| South West† | 0.13 | [0.06,0.33] | <0.001 | 1.56 | [0.94,2.59] | 0.08 | 0.73 | [0.50,1.07] | 0.10 | 0.95 | [0.68,1.34] | 0.78 | 0.95 | [0.68,1.32] | 0.76 |
| Intercept | 0.02 | [0.01,0.04] | <0.001 | 0.02 | [0.01,0.02] | <0.001 | 0.02 | [0.01,0.02] | <0.001 | 0.02 | [0.02,0.02] | <0.001 | 0.02 | [0.02,0.03] | <0.001 |
| Random estimates | | | | | | | | | | | | | | | |
| $\sigma_u^2$ (log odds) | 7.58 | [6.02,9.54] | | 5.84 | [5.20,6.56] | | 3.00 | [2.72,3.32] | | 2.52 | [2.30,2.76] | | 2.34 | [2.15,2.56] | |

Notes: † Reference region: London

Table S9 Results on service decline from multilevel logit models (year 2023) using multiple imputation with delta adjustment (MI=20)

| | 2023 $\delta_{rural}$=-0.2,$\delta_{IMD}$=-0.2 (n=266,193 N=1,616) | | | 2023 $\delta_{rural}$=0.2,$\delta_{IMD}$=0.2 (n=266,193 N=1,616) | | | 2023 $\delta_{rural}$=-0.2,$\delta_{IMD}$=0.2 (n=266,193 N=1,616) | | | 2023 $\delta_{rural}$=-0.5,$\delta_{IMD}$=-0.5 (n=266,193 N=1,616) | | | 2023 $\delta_{rural}$=0.5,$\delta_{IMD}$=0.5 (n=266,193 N=1,616) | | |
|---|---|---|---|---|---|---|---|---|---|---|---|---|---|---|---|
| | OR | 95% CI | p | OR | 95% CI | p | OR | 95% CI | p | OR | 95% CI | p | OR | 95% CI | p |
| Age: 30-39 (vs <30) | 1.24 | [1.14,1.35] | <0.001 | 1.25 | [1.14,1.36] | <0.001 | 1.23 | [1.13,1.34] | <0.001 | 1.23 | [1.13,1.34] | <0.001 | 1.24 | [1.14,1.35] | <0.001 |
| Age: 40-49 (vs <30) | 1.50 | [1.38,1.62] | <0.001 | 1.50 | [1.39,1.63] | <0.001 | 1.49 | [1.37,1.61] | <0.001 | 1.49 | [1.37,1.61] | <0.001 | 1.50 | [1.38,1.62] | <0.001 |
| Age: 50-59 (vs <30) | 1.81 | [1.68,1.96] | <0.001 | 1.82 | [1.69,1.97] | <0.001 | 1.81 | [1.68,1.95] | <0.001 | 1.81 | [1.67,1.95] | <0.001 | 1.81 | [1.68,1.96] | <0.001 |
| Age: 60-69 (vs <30) | 2.58 | [2.39,2.78] | <0.001 | 2.59 | [2.40,2.80] | <0.001 | 2.57 | [2.38,2.77] | <0.001 | 2.57 | [2.39,2.77] | <0.001 | 2.59 | [2.40,2.79] | <0.001 |
| Age: 70-79 (vs <30) | 3.11 | [2.89,3.36] | <0.001 | 3.13 | [2.90,3.38] | <0.001 | 3.10 | [2.87,3.34] | <0.001 | 3.10 | [2.88,3.34] | <0.001 | 3.11 | [2.89,3.36] | <0.001 |
| Age: 80+ (vs <30) | 2.70 | [2.50,2.91] | <0.001 | 2.71 | [2.51,2.92] | <0.001 | 2.68 | [2.48,2.89] | <0.001 | 2.69 | [2.49,2.90] | <0.001 | 2.69 | [2.50,2.90] | <0.001 |
| Female (vs male) | 0.79 | [0.77,0.82] | <0.001 | 0.79 | [0.77,0.82] | <0.001 | 0.79 | [0.77,0.82] | <0.001 | 0.79 | [0.77,0.82] | <0.001 | 0.79 | [0.77,0.82] | <0.001 |
| White (vs other) | 1.33 | [1.26,1.40] | <0.001 | 1.32 | [1.26,1.39] | <0.001 | 1.33 | [1.26,1.40] | <0.001 | 1.33 | [1.26,1.40] | <0.001 | 1.33 | [1.26,1.40] | <0.001 |
| IMD 2 (vs 1) | 1.00 | [0.92,1.09] | 0.93 | 1.00 | [0.92,1.09] | 0.95 | 0.98 | [0.90,1.07] | 0.72 | 1.04 | [0.95,1.13] | 0.37 | 0.99 | [0.91,1.07] | 0.75 |
| IMD 3 (vs 1) | 0.98 | [0.90,1.07] | 0.68 | 0.96 | [0.88,1.05] | 0.38 | 0.95 | [0.88,1.04] | 0.28 | 1.01 | [0.93,1.10] | 0.76 | 0.94 | [0.86,1.03] | 0.21 |
| IMD 4 (vs 1) | 1.05 | [0.97,1.14] | 0.23 | 1.03 | [0.95,1.12] | 0.47 | 1.02 | [0.94,1.11] | 0.66 | 1.09 | [1.00,1.18] | 0.05 | 1.00 | [0.92,1.09] | 0.91 |
| IMD 5 (vs 1) | 0.98 | [0.90,1.07] | 0.68 | 0.96 | [0.87,1.05] | 0.38 | 0.95 | [0.86,1.04] | 0.24 | 1.02 | [0.93,1.12] | 0.65 | 0.94 | [0.86,1.02] | 0.15 |
| IMD 6 (vs 1) | 1.03 | [0.95,1.12] | 0.45 | 1.01 | [0.92,1.10] | 0.87 | 1.00 | [0.92,1.09] | 0.96 | 1.08 | [0.99,1.18] | 0.08 | 0.98 | [0.90,1.07] | 0.72 |
| IMD 7 (vs 1) | 1.02 | [0.94,1.11] | 0.60 | 1.00 | [0.91,1.08] | 0.92 | 0.99 | [0.91,1.08] | 0.87 | 1.07 | [0.98,1.16] | 0.11 | 0.97 | [0.89,1.05] | 0.48 |
| IMD 8 (vs 1) | 1.07 | [0.98,1.17] | 0.11 | 1.04 | [0.96,1.14] | 0.34 | 1.04 | [0.95,1.14] | 0.39 | 1.11 | [1.02,1.21] | 0.01 | 1.02 | [0.94,1.12] | 0.60 |
| IMD 9 (vs 1) | 1.06 | [0.98,1.16] | 0.16 | 1.04 | [0.95,1.14] | 0.37 | 1.04 | [0.95,1.13] | 0.43 | 1.10 | [1.01,1.20] | 0.02 | 1.02 | [0.94,1.11] | 0.65 |
| IMD 10 (vs 1) | 0.98 | [0.90,1.08] | 0.75 | 0.99 | [0.90,1.08] | 0.81 | 0.98 | [0.90,1.08] | 0.72 | 1.00 | [0.91,1.10] | 0.94 | 0.99 | [0.91,1.08] | 0.83 |
| Rural (vs urban) | 1.07 | [0.99,1.15] | 0.08 | 1.05 | [0.97,1.14] | 0.20 | 1.06 | [0.98,1.15] | 0.12 | 1.07 | [0.99,1.16] | 0.10 | 1.03 | [0.96,1.11] | 0.40 |
| North East† | 1.99 | [1.31,3.03] | 0.001 | 1.99 | [1.31,3.03] | 0.001 | 1.99 | [1.30,3.03] | 0.001 | 1.99 | [1.31,3.03] | 0.001 | 1.98 | [1.30,3.01] | 0.001 |
| North West† | 1.51 | [1.18,1.93] | 0.001 | 1.51 | [1.18,1.92] | 0.001 | 1.50 | [1.17,1.91] | 0.001 | 1.52 | [1.19,1.93] | 0.001 | 1.50 | [1.18,1.91] | 0.001 |
| Yorkshire & Humber† | 1.53 | [0.86,2.72] | 0.15 | 1.53 | [0.86,2.73] | 0.15 | 1.52 | [0.85,2.71] | 0.16 | 1.54 | [0.86,2.74] | 0.15 | 1.52 | [0.86,2.72] | 0.15 |
| East Midlands† | 1.07 | [0.59,1.92] | 0.83 | 1.07 | [0.60,1.94] | 0.81 | 1.04 | [0.58,1.88] | 0.89 | 1.07 | [0.59,1.92] | 0.83 | 1.07 | [0.59,1.92] | 0.83 |
| West Midlands† | 1.42 | [1.11,1.82] | 0.005 | 1.40 | [1.10,1.80] | 0.007 | 1.41 | [1.10,1.80] | 0.007 | 1.42 | [1.11,1.82] | 0.005 | 1.42 | [1.11,1.81] | 0.005 |
| East of England† | 1.03 | [0.63,1.69] | 0.90 | 1.01 | [0.62,1.66] | 0.96 | 1.02 | [0.62,1.68] | 0.93 | 1.04 | [0.63,1.70] | 0.88 | 1.01 | [0.62,1.66] | 0.97 |
| South East† | 0.99 | [0.76,1.28] | 0.92 | 0.98 | [0.76,1.28] | 0.90 | 0.97 | [0.75,1.27] | 0.85 | 0.99 | [0.76,1.28] | 0.93 | 0.98 | [0.76,1.28] | 0.90 |
| South West† | 0.95 | [0.69,1.32] | 0.77 | 0.95 | [0.69,1.32] | 0.77 | 0.95 | [0.69,1.32] | 0.76 | 0.95 | [0.68,1.31] | 0.74 | 0.95 | [0.69,1.32] | 0.78 |
| Intercept | 0.02 | [0.02,0.03] | <0.001 | 0.02 | [0.02,0.03] | <0.001 | 0.02 | [0.02,0.03] | <0.001 | 0.02 | [0.02,0.03] | <0.001 | 0.02 | [0.02,0.03] | <0.001 |
| Random estimates | | | | | | | | | | | | | | | |
| $\sigma_u^2$ (log odds) | 2.34 | [2.14,2.55] | | 2.35 | [2.15,2.56] | | 2.35 | [2.15,2.56] | | 2.34 | [2.15,2.56] | | 2.33 | [2.14,2.55] | |

Notes: † Reference region: London

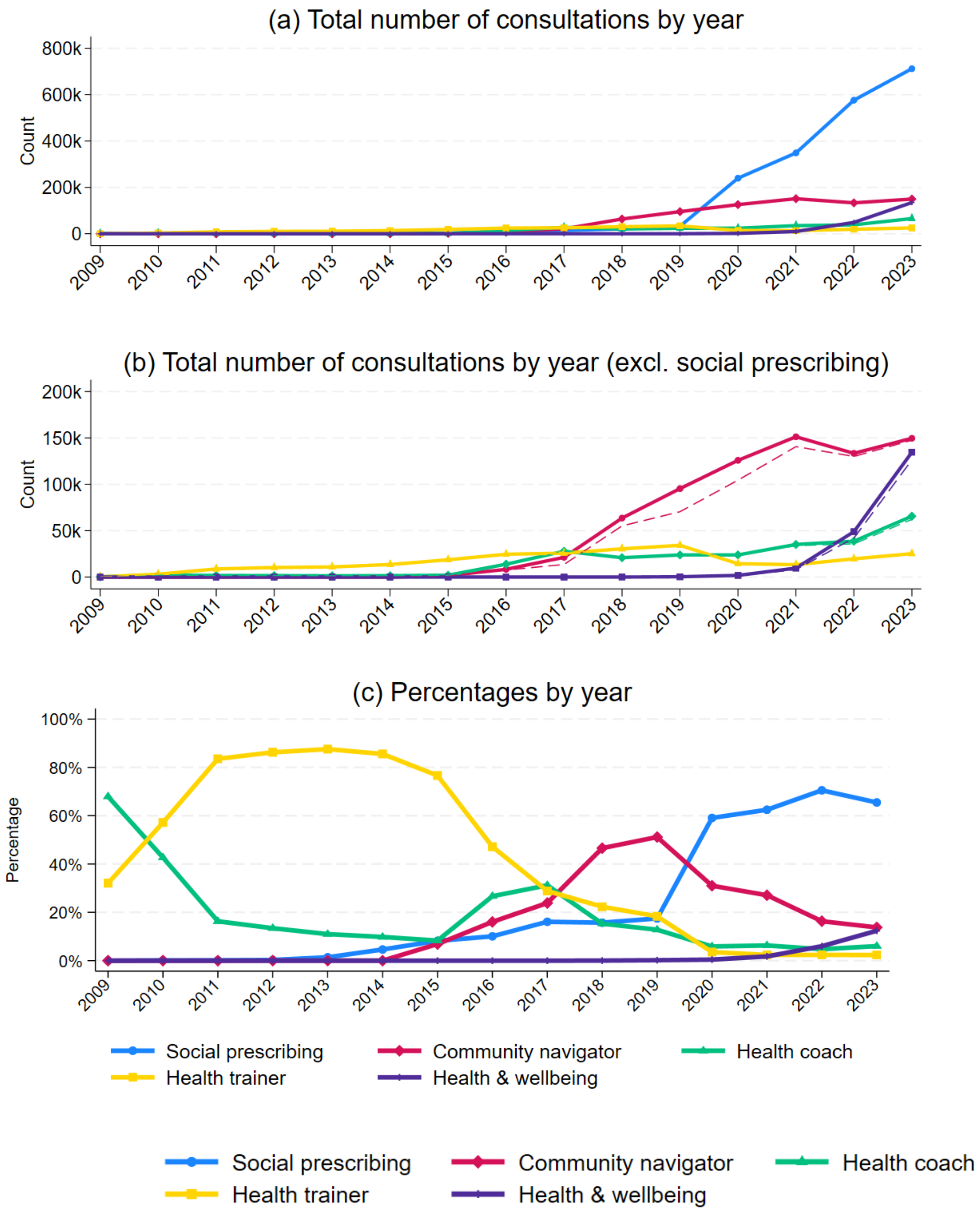

Figure S1. Numbers and percentages of GP consultations related to social prescribing and other non-clinical interventions by year. Notes: dashed lines in Figure 1b removing double counts if other non-clinical codes were used in with social prescribing in the same consultation

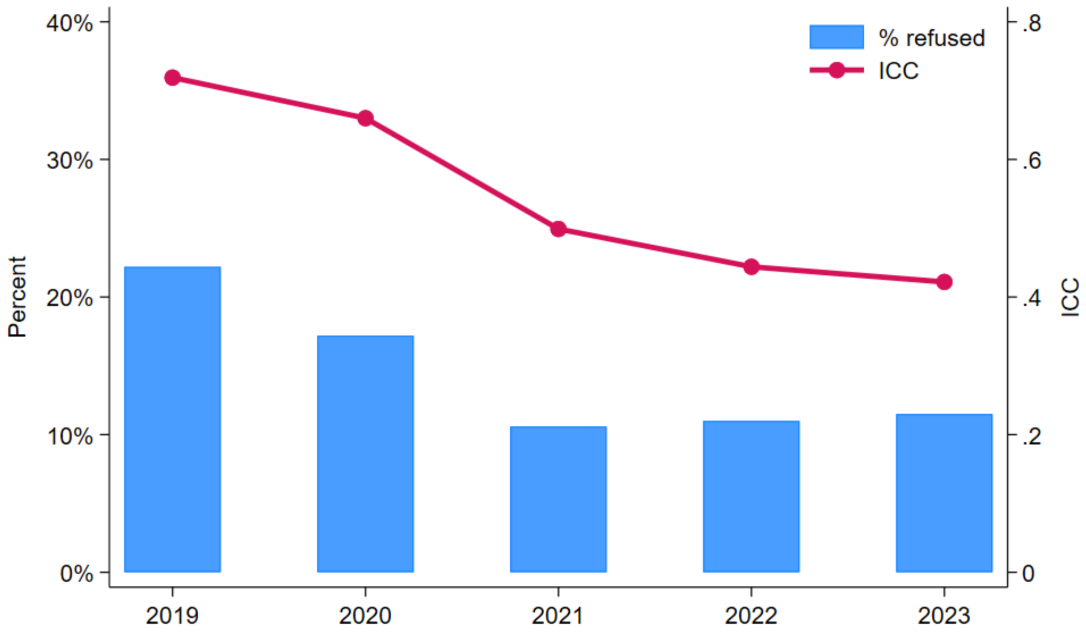

Figure S2. Refusal rates by year and intraclass correlation coefficients (ICCs) from unconditional multilevel logistic regression models by year
